# Relative perceived importance of different settings for SARS-CoV2 acquisition in England and Wales: Analysis of the Virus Watch Community Cohort

**DOI:** 10.1101/2021.06.11.21258730

**Authors:** Sarah Beale, Thomas Byrne, Ellen Fragaszy, Jana Kovar, Vincent Nguyen, Anna Aryee, Wing Lam Erica Fong, Cyril Geismar, Parth Patel, Madhumita Shrotri, Nicholas Patni, Isobel Braithwaite, Annalan M D Navaratnam, Robert W Aldridge, Andrew Hayward

## Abstract

We aimed to assess the relative importance of different settings for SARS-CoV2 transmission in a large community cohort. We demonstrate the importance of home, work and education as venues for transmission. In children, education was most important and in older adults essential shopping was of high importance. Our findings support public health messaging about infection control at home, advice on working from home and restrictions in different venues.

## Background

Risk factors for the transmission of SARS-CoV2 are understood to be driven by the complex interplay of a range of factors that can be broadly classified into four main areas[1] including viral load dynamics[2], contact patterns[3,4], environmental factors[5] and socioeconomic inequalities[6–8]. However, the relative contribution of different settings to overall transmission has proven difficult to quantify at a population level.[1] A lack of evidence about their relative contribution to transmission limits our ability to make evidence-based decisions about which settings should be the focus of control measures at different stages of the pandemic as different public health and policy actions are required. In this analysis, we aimed to assess the perceived relative importance of different settings for SARS-CoV2 transmission in a large community cohort (Virus Watch).

## Perceived Setting of SARS-CoV-2 Acquisition

Virus Watch is a household community cohort study which began recruitment in June 2020 and has recruited circa 50,000 individuals across England and Wales with weekly online follow up and self-reporting of any positive SARS-CoV2 virological tests.[9] On the 17th of March 2021, participants were asked, “If you have had a positive COVID-19 swab test at any point in the pandemic, did you test positive after:…”. Possible responses were “Contact with confirmed COVID-19 case(s)”, “Contact with suspected COVID-19 case(s)”, “No known contact with confirmed or suspected COVID-19 case(s)”, or “I have not had a positive COVID-19 swab test during the pandemic”. Participants who answered anything other than “I have not had a positive COVID-19 swab test during the past month” were then asked “Where do you think you may have caught COVID-19?”. Participants were allowed to select more than one setting category and we classified these settings into the following groups: home, someone else’s home, work, place of education, public transport, essential shop, healthcare setting, leisure (comprising ‘bar, pub, or club’, ‘eating out in a restaurant, cafe, or canteen’, ‘gym/indoor sports facility’, ‘hairdresser, barber, nail salon, or similar location’, and ‘shop for non-essential items’), and other. Survey responses were linked to a previously reported date of infection, where available.

Using these data, we described the perceived setting of SARS-CoV-2 acquisition by contact status and stratified these descriptive analyses by age of the case and by time period of infection (Jan-May 2020, Jun-Aug 2020, Sep 2020-Dec 2020, Jan-March 2021). The survey was sent to 21,444 households comprising 45,654 study participants on 17/03/2021, and 18,096 (40%) individual participants had completed the survey in full when data were extracted on 28/03/2021. Of these, 1142 participants had self-reported a positive SARS-CoV2 test previously during follow-up and responded to the main question of interest. 499 (44%) reported known contact with a confirmed case, 120 (10%) contact with a suspected case and 523 (46%) reported no known contact. Amongst all cases, the perceived setting of acquisition was, in descending order of frequency, within the home (n=317, 33%), at work (n=259, 23%), in an essential shop (n=201, 18%), other venues (n=129, 11%), in a leisure venue (n=112, 10%), in a place of education (n=96, 8%), in healthcare settings (n=90, 8%), on public transport (n=75, 7%), and in someone else’s home (n=64, 6%; Table 1). This varied considerably by whether or not the person was a contact of a known case or not (Figure 1).

**Table 1.**
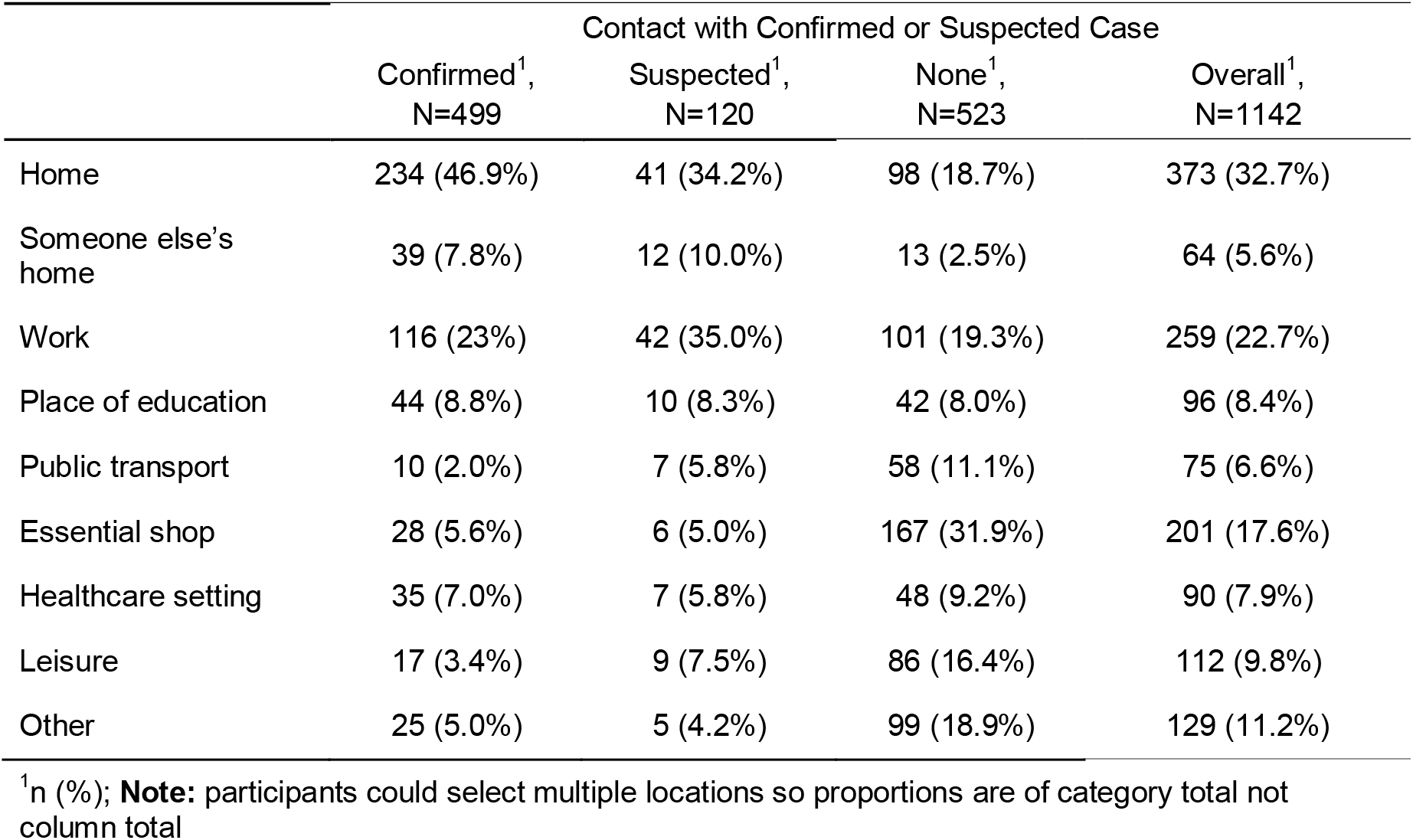
Perceived Setting of SARS-CoV2 Acquisition by Contact Status I.

|  | Contact with Confirmed or Suspected Case |  |  | Overall<sup>1</sup>,<br>N=1142 |
| --- | --- | --- | --- | --- |
|  | Confirmed<sup>1</sup>,<br>N=499 | Suspected<sup>1</sup>,<br>N=120 | None<sup>1</sup>,<br>N=523 |  |
| Home | 234 (46.9%) | 41 (34.2%) | 98 (18.7%) | 373 (32.7%) |
| Someone else's home | 39 (7.8%) | 12 (10.0%) | 13 (2.5%) | 64 (5.6%) |
| Work | 116 (23%) | 42 (35.0%) | 101 (19.3%) | 259 (22.7%) |
| Place of education | 44 (8.8%) | 10 (8.3%) | 42 (8.0%) | 96 (8.4%) |
| Public transport | 10 (2.0%) | 7 (5.8%) | 58 (11.1%) | 75 (6.6%) |
| Essential shop | 28 (5.6%) | 6 (5.0%) | 167 (31.9%) | 201 (17.6%) |
| Healthcare setting | 35 (7.0%) | 7 (5.8%) | 48 (9.2%) | 90 (7.9%) |
| Leisure | 17 (3.4%) | 9 (7.5%) | 86 (16.4%) | 112 (9.8%) |
| Other | 25 (5.0%) | 5 (4.2%) | 99 (18.9%) | 129 (11.2%) |

<sup>1</sup>n (%); **Note:** participants could select multiple locations so proportions are of category total not column total

**Figure 1.**
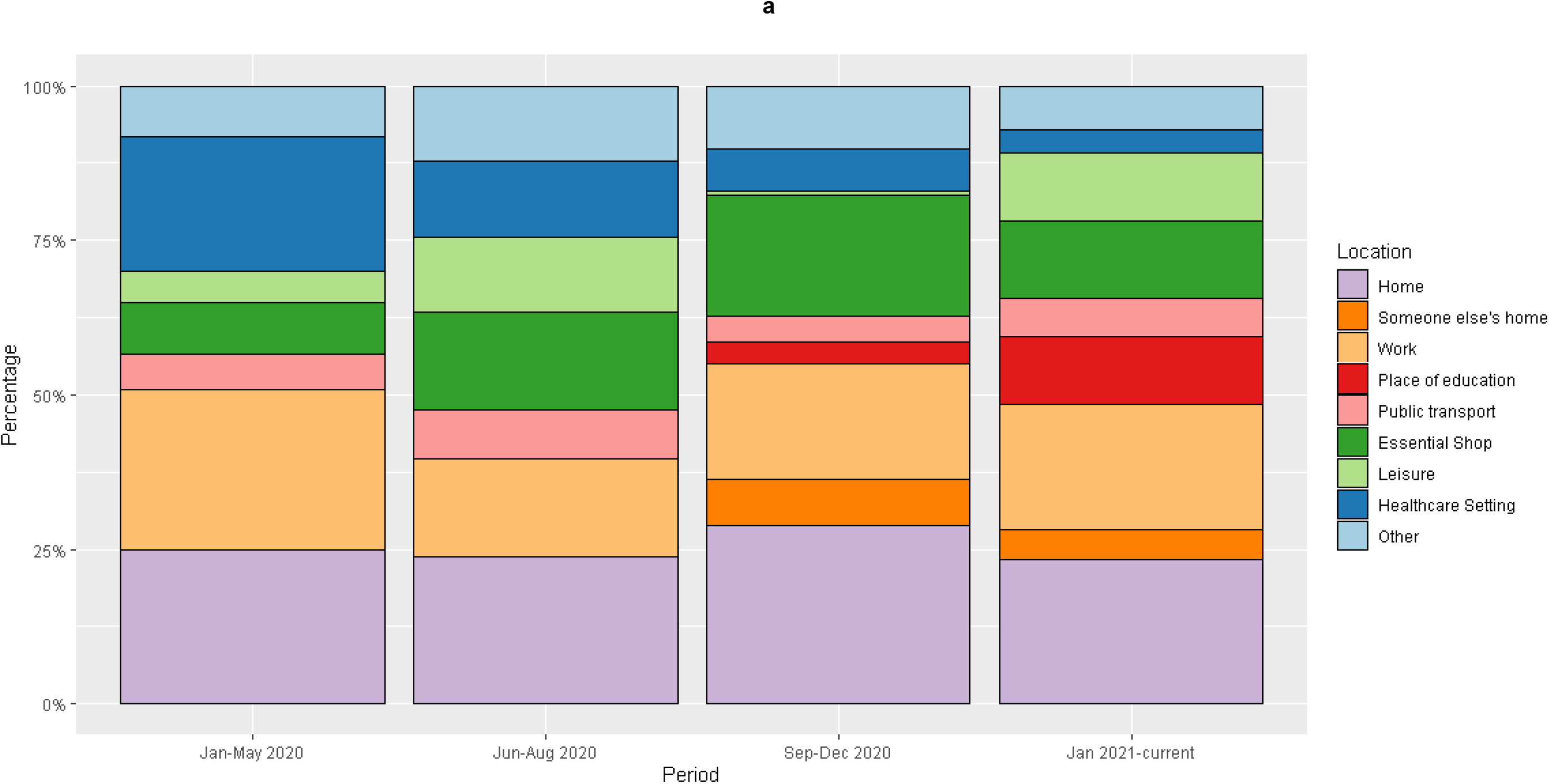
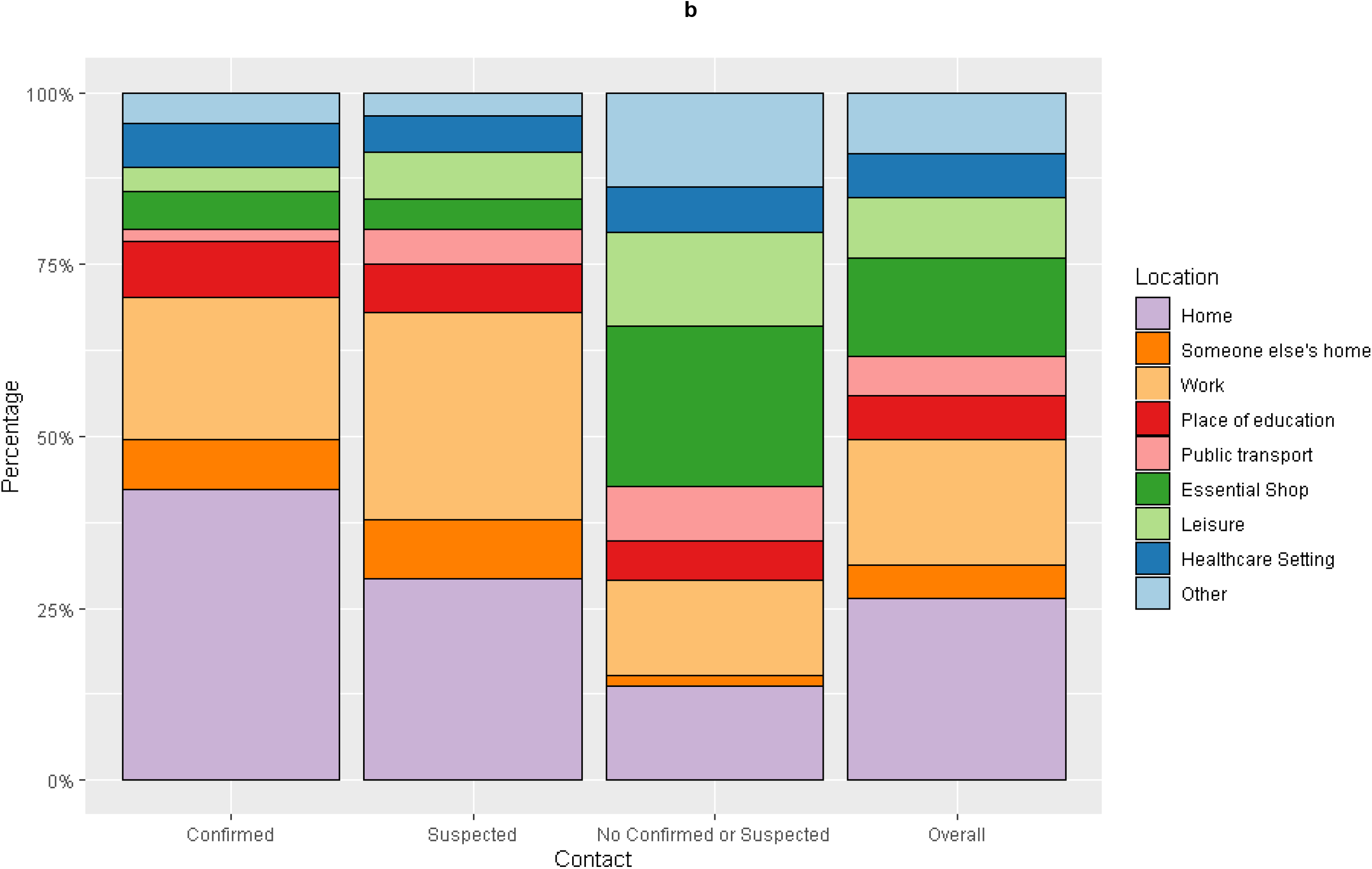
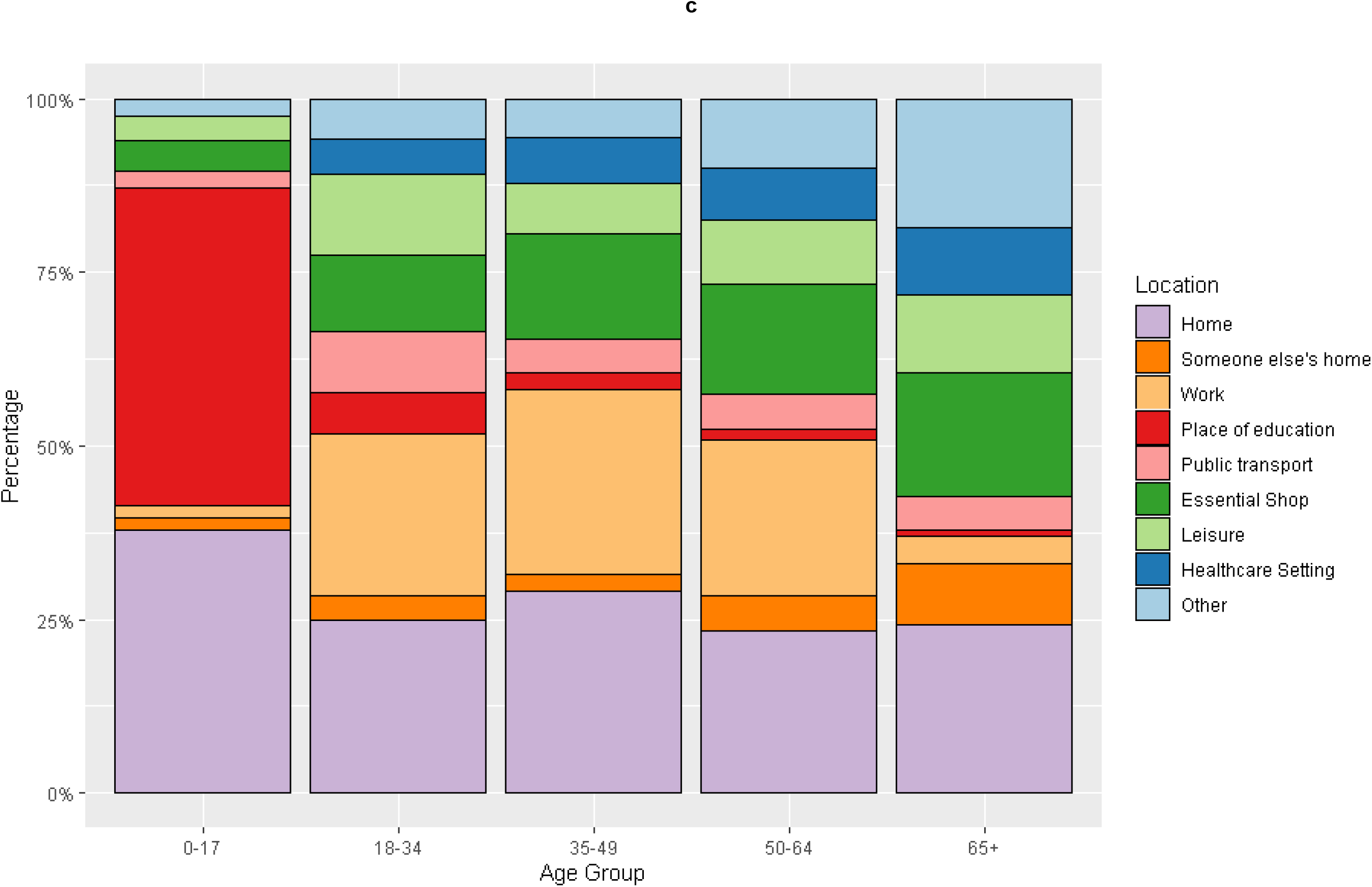
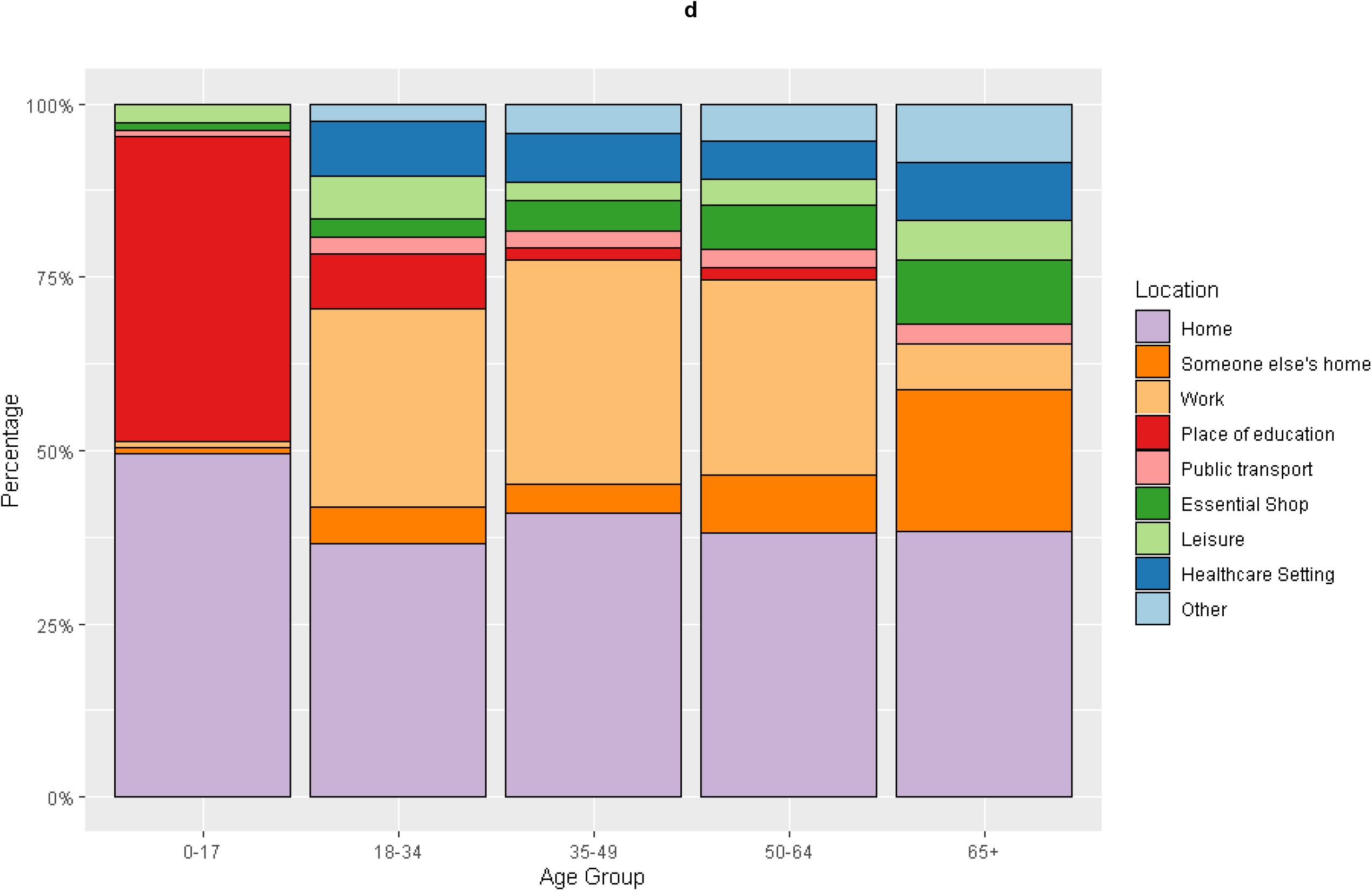
Perceived Setting of SARS-CoV2 Acquisition by (a) Time Period, (b) Contact Status, (c) Age and (d) Age – Contacts of Confirmed or Suspected Cases. **Note:** Participants could select multiple locations so proportions are calculated from group *n* and may sum to >100.0%

For those with known contact with a confirmed case, the perceived venues for transmission in descending order of frequency were: home (n=234, 47%), work (n=116, 23%), education (n=44, 9%), someone else’s home (n=39, 8%), healthcare setting (n=35, 7%), essential shop (n=28, 6%), other venues (n=25, 5%), and leisure venue (n=17, 3%). For those with no recognized contact, the perceived venues for acquisition were: an essential shop (n=167, 32%), work (n=101, 19%), other venues (n=99, 19%), home (n=98, 19%), a leisure venue (n=86, 16%), transport (n=58, 11%), healthcare (n=48, 9%), education (n=42, 8%), and someone else’s home (n=13, 3%).

The most important perceived venue for transmission varied by age group: place of education was more important than home for children aged 0-17, home and workplace were of similar importance for working age adults, and home and essential shops were the most important settings for those aged over 65 years (Table 2). Young adults aged 18-34 were more likely than other age groups to perceive leisure venues and public transport as the venue of acquisition.

**Table 2.** Perceived Setting of SARS-CoV2 Acquisition by Age*.

|  | Overall |  |  |  |  | Contact with Confirmed or Suspected Case |  |  |  |  |
| --- | --- | --- | --- | --- | --- | --- | --- | --- | --- | --- |
|  | 0 to 17,<br>N = 120<sup>1</sup> | 18 to 34,<br>N = 191<sup>1</sup> | 35 to 49,<br>N = 262<sup>1</sup> | 50 to 64,<br>N = 361<sup>1</sup> | 65+,<br>N = 206<sup>1</sup> | 0 to 17,<br>N = 78<sup>1</sup> | 18 to 34,<br>N = 106<sup>1</sup> | 35 to 49,<br>N = 152<sup>1</sup> | 50 to 64,<br>N = 194<sup>1</sup> | 65+,<br>N = 87<sup>1</sup> |
| Home | 53<br>(44.2%) | 64<br>(33.5%) | 94<br>(35.9%) | 100<br>(27.7%) | 61<br>(29.6%) | 41<br>(52.6%) | 45<br>(42.4%) | 71<br>(46.7%) | 81<br>(41.7%) | 36<br>(41.4%) |
| Someone else's home | 2 (1.7%) | 9 (4.7%) | 9 (3.4%) | 21<br>(5.8%) | 23<br>(11.2%) | 1 (1.3%) | 6 (5.7%) | 7 (4.6%) | 18 (9.3%) | 19<br>(21.8%) |
| Work | 2 (1.7%) | 61<br>(31.9%) | 87<br>(33.2%) | 98<br>(27.1%) | 11<br>(5.3%) | 1 (1.3%) | 35<br>(33.0%) | 56<br>(36.8%) | 60<br>(30.9%) | 6 (6.9%) |
| Place of education | 63<br>(52.5%) | 15 (7.9%) | 8 (3.1%) | 8 (2.2%) | 1 (0.5%) | 37<br>(47.4%) | 10 (9.4%) | 3 (2.0%) | 3 (1.5%) | 0 (0.0%) |
| Public transport | 4 (3.3%) | 22<br>(11.5%) | 15 (5.7%) | 22<br>(6.1%) | 12<br>(5.8%) | 1 (1.3%) | 3 (2.8%) | 4 (2.6%) | 6 (3.1%) | 3 (3.4%) |
| Essential shop | 6 (5.0%) | 28<br>(14.7%) | 51<br>(19.5%) | 70<br>(19.4%) | 46<br>(22.3%) | 1 (1.3%) | 3 (2.8%) | 7 (4.6%) | 14 (7.2%) | 9<br>(10.3%) |
| Healthcare setting | 0 (0.0%) | 14 (7.3%) | 20 (7.6%) | 32<br>(8.9%) | 24<br>(11.7%) | 0 (0.0%) | 10 (9.4%) | 12 (7.9%) | 12 (6.2%) | 8 (9.2%) |
| Leisure | 5 (4.2%) | 28<br>(14.7%) | 19 (7.3%) | 36<br>(10.0%) | 24<br>(11.7%) | 2 (2.6%) | 7 (6.6%) | 4 (2.6%) | 8 (4.1%) | 5 (5.7%) |
| Other | 4 (3.3%) | 15 (7.9%) | 19 (7.3%) | 43<br>(11.9%) | 48<br>(23.3%) | 0 (0.0%) | 3 (2.8%) | 7 (4.6%) | 12 (6.2%) | 8 (9.2%) |

<sup>1</sup>n (%); \* Two cases excluded due to missing age; **Note:** Participants could select multiple locations so proportions are calculated from group *n* and may sum to >100.0%

Home settings had a relatively stable level of perceived importance between Jan-May 2020 (n=21, 34%) and Jan-March 2021 (n=106, 30%; Table 3). The proportions for someone else’s home increased over time (n=0, 0% Jan-May 2020 vs. n=27, 9% Jan-March 2021), as did place of education (n=0, 0% vs. n=12, 4%). The proportion who perceived that they were infected in a healthcare setting decreased over time (n=18, 26% vs. n=25, 8%). Caution should be used interpreting these time period changes because of small numbers for some groups, particularly in the Jun-Aug time period when levels of infection were low.

**Table 3.** Perceived Setting of SARS-CoV2 Acquisition by Time Period.

| Characteristic | Jan-May 2020, N = 70<sup>1</sup> | Jun-Aug 2020, N = 18<sup>1</sup> | Sep 2020-Dec 2020, N = 377<sup>1</sup> | Jan 2021-March 2021, N = 311<sup>1</sup> |
| --- | --- | --- | --- | --- |
| Home | 21 (30.0%) | 6 (33.3%) | 113 (30.0%) | 106 (34.1%) |
| Someone else's home | 0 (0.0%) | 0 (0.0%) | 22 (5.8%) | 27 (8.7%) |
| Work | 22 (31.4%) | 4 (22.2%) | 97 (25.7%) | 69 (22.2%) |
| Place of education | 0 (0.0%) | 0 (0.0%) | 54 (14.3%) | 12 (3.9%) |
| Public transport | 5 (7.1%) | 2 (11.1%) | 30 (8.0%) | 16 (5.1%) |
| Essential shop | 7 (10.0%) | 4 (22.2%) | 62 (16.4%) | 71 (22.8%) |
| Healthcare setting | 18 (25.7%) | 3 (16.7%) | 17 (4.5%) | 25 (8.0%) |
| Leisure | 4 (5.7%) | 3 (16.7%) | 53 (14.1%) | 4 (1.3%) |
| Other | 7 (10.0%) | 3 (16.7%) | 34 (9.0%) | 37 (11.9%) |

<sup>1</sup>n (%); **Note:** Participants could select multiple locations so proportions are calculated from group *n* and may sum to >100.0%

## Discussion

Our findings illustrate the central perceived importance of home, work and place of education as venues for transmission, although the relative importance of different settings is likely to change over time and with variation in restrictions. In children, place of education is most important, and in older adults essential shopping is of high importance. Our estimates cover several periods of intense restrictions. As these restrictions loosen, the relative importance of out of household transmission is likely to increase.

Our study was reliant on community testing for case ascertainment so are most likely to represent infections acquired after the first UK lockdown following initiation of the Test, Trace and Isolate programme. Perceived venues of acquisition in those with known contact are likely biased toward venues such as home, work and education where contact tracing can readily lead to identification of known contact. Conversely those with no known contact may be more likely to conclude they acquired SARS-CoV2 infection in settings where they are in contact with strangers (such as in shops).

Considering these possible biases in reporting of the importance of these perceived settings by known or unknown contact status, our estimates provide useful upper and lower ranges for the likely relative importance of acquisition of SARS-CoV2 in England and Wales over our study period. In future waves of transmission, these findings can support public health messaging about infection control in the home, advice on working from home, restrictions in different venues, and advice to vulnerable elderly to reduce exposure to shops, for example through online shopping.

## Data Availability

We aim to share aggregate data from this project on our website and via a "Findings so far" section on our website - https://ucl-virus-watch.net/. We will also be sharing individual record-level data on a research data-sharing service such as the Office of National Statistics Secure Research Service. In sharing the data we will work within the principles set out in the UKRI Guidance on best practice in the management of research data. Access to use of the data whilst research is being conducted will be managed by the Chief Investigators (ACH and RWA) in accordance with the principles set out in the UKRI guidance on best practice in the management of research data. We will put analysis code on publicly available repositories to enable their reuse.

